# Clinical, Radiological Profile, Risk Factors, and Short-Term Outcomes of Meconium Aspiration Syndrome Among Neonates at Two Referral Hospitals in Uganda

**DOI:** 10.1101/2025.11.17.25340437

**Authors:** Khalif Guled Hersi, Kyomugisa Beatrice, Laker Gorety, Martin Nduwimana, Bappah Alkali, Abdikheyr Mohamed Aden, Ahmed khalif shire, Yasin Ahmed H. Abshir, Abdulahi Abdirizak Farah, Hamdi M. Yusuf, Theoneste Hakizimana, Bahari Yusuf, Walyeldin Elfakey, Grace Ndeezi

## Abstract

Meconium Aspiration Syndrome (MAS) is a significant contributor to neonatal morbidity and mortality, particularly in low-resource settings. Despite its clinical relevance, limited local data exist on the burden of MAS, clinical and radiological profile, associated risk factors, and short-term outcomes in Uganda. This study aimed to describe the clinical and radiological features, determine the proportion of neonates with MAS and its risk factors among neonates admitted to two regional referral hospitals in Uganda. A prospective cohort study was conducted among 125 neonates at Mubende and Fort Portal Regional Referral hospitals between June and August 2025. Neonates were consecutively enrolled and followed for up to 14 days of life. Data was collected using a structured tool covering clinical features, radiological findings, maternal and perinatal factors, and outcomes. Descriptive statistics summarized baseline characteristics and outcome proportions, while multivariate Poisson regression identified factors independently associated with MAS. Adjusted Incidence Rate Ratios (aIRR) and 95% Confidence Intervals (CI) were reported. Clinically, neonates most frequently presented with nasal flaring 23 (18.4%), tachypnoea 21(16.8%), chest retractions 19(15.2%), cyanosis 18(14.4%), and grunting 12(9.6%), wheezing 13 (10.4%), crackles 11 (8.8%). Radiological features included hyperinflation 6(4.8%), bilateral infiltrates 5(4.0%), and atelectasis 4(3.2%). Independent predictors of MAS included maternal age ≥35 years (aIRR = 2.09), pregnancy-induced diabetes mellitus (aIRR = 2.77, p = 0.04), fetal distress (aIRR = 3.97, p < 0.001), and a low 5th minute APGAR score of 0–3 (aIRR = 1.94, p = 0.03). In-hospital deaths (P = 0.002) and length of hospital stay (P < 0.001) were significantly associated with MAS among neonates in the study area. MAS remains a critical neonatal condition with a significant risk of mortality and prolonged hospitalization in Uganda. Advanced maternal age, gestational diabetes, fetal distress, and low APGAR scores were key predictors. Strengthening antenatal surveillance, improving intrapartum monitoring, and ensuring early neonatal resuscitation are essential to reduce the burden of MAS in resource-limited settings.

## Introduction

Meconium Aspiration Syndrome (MAS) is a major cause of neonatal respiratory distress and mortality, particularly in term and post-term infants born through meconium-stained amniotic fluid (MSAF). Globally, MSAF occurs in 5–25% of deliveries, and approximately 10% of these neonates develop MAS, with the risk increasing significantly in post-term pregnancies and in the presence of fetal-maternal stressors such as hypoxia and infection [1]. In Sub-Saharan Africa, MAS contributes to 5–12% of neonatal deaths, with Uganda reporting neonatal mortality rates as high as 27 deaths per 1,000 live births [2].

Clinically, MAS presents with respiratory distress within minutes to hours after birth, characterized by tachypnoea, cyanosis, nasal flaring, and chest retractions [3]. Radiologically, chest X-rays often reveal hyperinflation, bilateral patchy infiltrates, and atelectasis hallmarks of chemical pneumonitis and mechanical airway obstruction [4]. The severity of MAS ranges from mild to life-threatening, with complications such as persistent pulmonary hypertension of the newborn (PPHN), hypoxic-ischemic encephalopathy, and prolonged hospitalization [5].

The conceptual framework guiding this study is the Web of Causation model, which posits that MAS arises from a complex interplay of maternal, fetal, and intrapartum factors rather than a single direct cause [6]. Risk factors include advanced maternal age, gestational diabetes, fetal distress, low APGAR scores, and placental insufficiency [7, 8]. Understanding these interrelated determinants is essential for designing effective interventions and improving neonatal outcomes.

Despite the clinical burden of MAS in Uganda, there is limited local data describing its presentation, associated risk factors, and short-term outcomes. Most existing studies focus on MSAF rather than confirmed MAS cases and offer limited follow-up durations [9]. This knowledge gap impedes the development of evidence-based protocols tailored to resource-limited settings.

This study was conducted to address this gap by assessing the clinical and radiological profile, identifying key risk factors, and evaluating short-term outcomes of MAS among neonates admitted to Fort Portal and Mubende Regional Referral Hospitals in Uganda.

## Methods and materials

### Study Design and Population

This study utilized a prospective cohort design to assess the clinical and radiological profile, risk factors, and short-term outcomes of MAS among neonates admitted to Fort Portal and Mubende Regional Referral Hospitals in Uganda. The study was conducted between July and September 2025, targeting neonates born through meconium-stained amniotic fluid (MSAF) and admitted with clinical suspicion or confirmation of MAS.

A total of 125 neonates were consecutively enrolled and followed for up to 14 days post-admission. The study population included neonates delivered at or after term gestation, presenting with respiratory distress and radiological features consistent with MAS. Inclusion criteria focused on neonates with documented MSAF and clinical signs of MAS, while those with congenital anomalies or alternative diagnoses were excluded. These hospitals were selected due to their high neonatal admission rates and their role as regional referral centres serving diverse populations in Western and Central Uganda.

### Sample Size Determination

The sample size was calculated using Lesli and Kish’s formula:

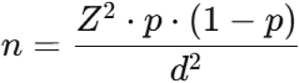

Where:

- n = required sample size
- Z standard normal deviation corresponding to the desired confidence level (1.96 at 95% confidence level)
- p = estimated prevalence of meconium aspiration syndrome (MAS) among neonates. Using findings by Okao at Kawempe National Referral Hospital, p=23.1% [9].
- d = margin of error (±5%)

A 95% confidence level (Z=1.96) was chosen to reduce sampling error, and a 5% margin of error (d = 0.05) was used, which is acceptable in epidemiological studies.

By substitution, n=114 study participants. Adding 10% to cater for loss of follow-up, the sample size required was 125

### Sampling Technique

This study employed a consecutive sampling technique to recruit eligible neonates born through meconium-stained amniotic fluid and admitted to the neonatal units of Fort Portal and Mubende Regional Referral Hospitals. All neonates who met the inclusion criteria during the study period from June to August 2025 were enrolled in the study as they presented, until the desired sample size was achieved.

### Study Procedure

Neonates born to mothers with meconium-stained amniotic fluid were clinically assessed for signs of respiratory distress, including tachypnoea, nasal flaring, chest retractions, cyanosis, grunting, wheezing, and crackles. A chest X-ray was performed to confirm the diagnosis of Meconium Aspiration Syndrome, with radiological features such as hyperinflation, bilateral infiltrates, and atelectasis considered diagnostic. A detailed maternal and neonatal history, along with a physical examination, was conducted to establish the clinical profile of each participant.

Participants were evaluated daily until discharge to monitor for short-term outcomes. Clinical assessments included documentation of respiratory status, episodes of apnoea, and any deterioration in condition. Information was gathered through bedside evaluations, maternal interviews, and consultations with ICU nursing staff. Neonates requiring mechanical ventilation were referred to Mulago National Referral Hospital for advanced care. At discharge, a final clinical evaluation was conducted to assess for residual respiratory distress. All enrolled neonates were prospectively followed from admission until the 14th day of life. Daily follow-up included review of treatment charts, nursing notes, and laboratory and radiological investigations. The primary outcomes assessed were in-hospital mortality and duration of hospital stay. Neonates discharged before day 14 were followed until discharge, while those still admitted on day 14 were censored at that point and their outcomes recorded.

### Data Quality Control

To ensure the integrity and reliability of the data collected, several quality control measures were implemented throughout the study. All research assistants underwent training on the study protocol, data collection tools, and ethical conduct prior to the commencement of data collection. The principal investigator supervised the data collection process daily, conducted spot checks, and reviewed completed forms for accuracy and completeness.

Standardized data collection instruments were used to minimize variability and ensure consistency across study sites. Daily ward rounds were conducted jointly by the principal investigator and clinical staff to verify clinical findings and ensure uniform documentation. Any discrepancies or missing data were promptly addressed through consultation with attending clinicians and review of patient records.

Double data entry was performed to reduce transcription errors, and the final dataset was cross-checked against original forms before analysis. Regular meetings were held with the research team to discuss challenges encountered and implement corrective actions. These measures ensured that the data collected were valid, reliable, and suitable for robust statistical analysis.

### Data Management and Analysis

All collected data were reviewed daily for completeness and accuracy by the principal investigator and trained research assistants. Data were entered into a secure, password-protected electronic database using double-entry verification to minimize transcription errors. The database was regularly backed up and access was restricted to authorized study personnel to ensure confidentiality and data integrity.

Data were analysed using STATA version 15. Descriptive statistics were used to summarize baseline characteristics, clinical and radiological profiles. The variables were presented as frequencies and percentages. To identify risk factors associated with Meconium Aspiration Syndrome (MAS) bivariate analysis was initially performed using modified Poisson regression analysis. Variables with a p-value <0.2 in bivariate analysis were included in a multivariate Poisson regression model to determine independent predictors of MAS and adverse outcomes. Adjusted Incidence Rate Ratios (aIRR) with 95% Confidence Intervals (CI) were reported, and statistical significance was set at p < 0.05. Fisher’s exact test was done to assess the association between MAS and short-term outcomes.

### Ethical Consideration

Ethical approval was obtained from the Kampala International University Research and Ethics Committee with REC number 2025-921. Informed consent was obtained from parents of all participants after explaining the purpose and procedures of the study. Confidentiality was maintained by anonymizing data, and participation was entirely voluntary, with the right to withdraw at any time without consequence.

## Results

### Socio-demographic characteristics of study participants

The majority of the mothers for the neonates (45.6%, 57/125) were aged 20-34 years, with primary education as their maximum level attained (37.6%, 47/125), unemployed (45.6%, 57/125), and residing in rural areas (52.8%, 66/125) (table 1).

**Table 1:** Sociodemographic characteristics of study participants.

| Variable | Frequencies (N=125) | Percentages (%) |
| --- | --- | --- |
| Maternal age (years) |  |  |
| <20 | 25 | 20.0 |
| 20-34 | 57 | 45.6 |
| 35+ | 43 | 34.4 |
| Mother's education level |  |  |
| No education | 33 | 26.4 |
| Primary | 47 | 37.6 |
| Secondary | 45 | 36.0 |
| Occupation |  |  |
| Employed | 68 | 54.4 |
| Unemployed | 57 | 45.6 |
| Residence type |  |  |
| Urban | 59 | 47.2 |
| Rural | 66 | 52.8 |
| Sex of child |  |  |
| Male | 67 | 53.6 |
| Female | 58 | 46.4 |

### Maternal characteristics of study participants

The majority of the mothers (64.8%, 81/125) reported having suffered from bacterial infection during pregnancy, and 12.8% (16/125) had pregnancy-induced hypertension. Additionally, 41.6% (52/125) had experienced obstetric haemorrhage during pregnancy, and 14.4% (18/125) had pregnancy-induced diabetes mellitus. A smaller proportion, 13.6% (17/125), had a history of PROM, and 65.6% (82/125) had spontaneous vaginal delivery. Additionally, 11.2% (14/125) had twins (Table 2).

**Table 2:**
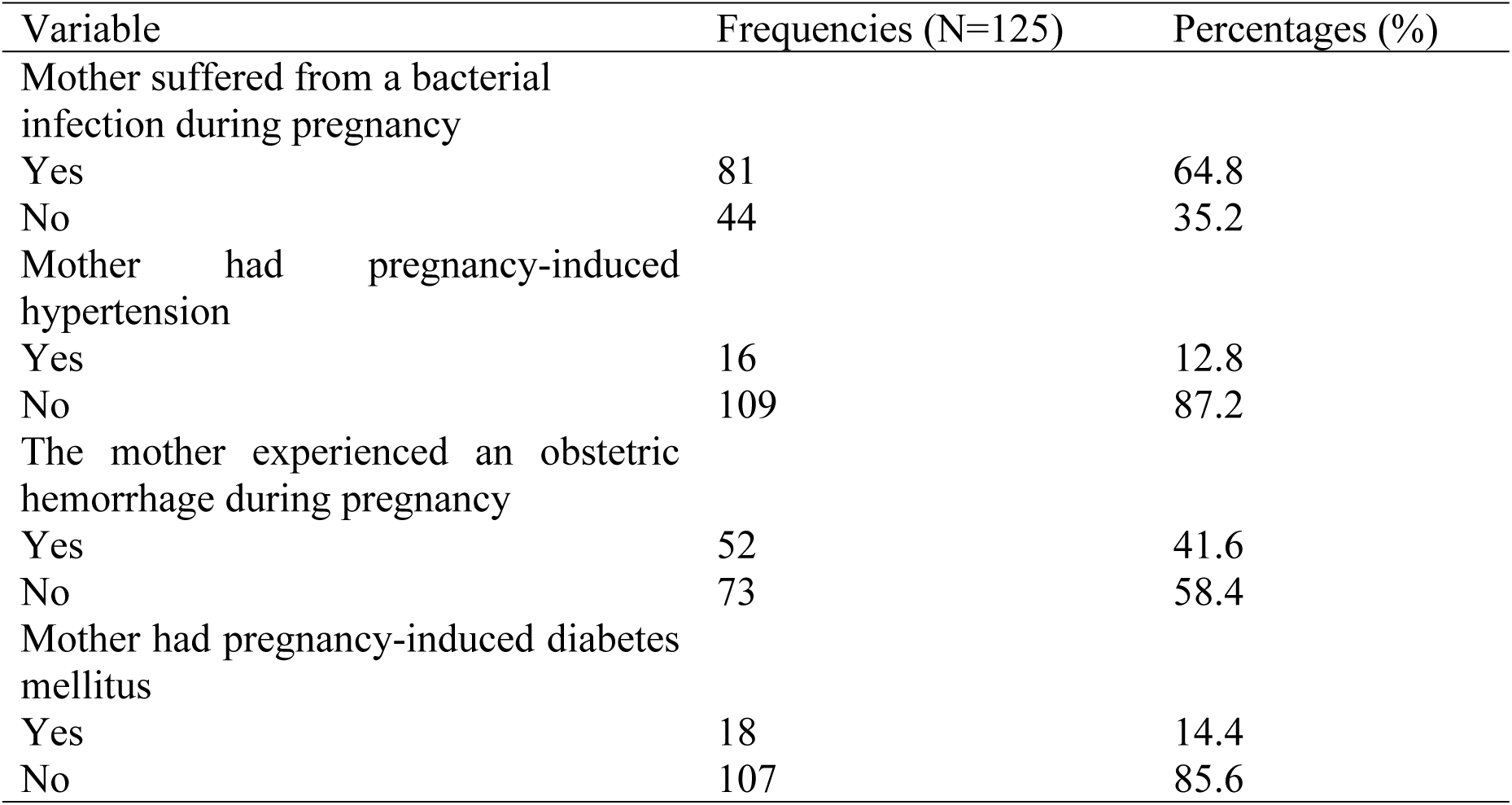
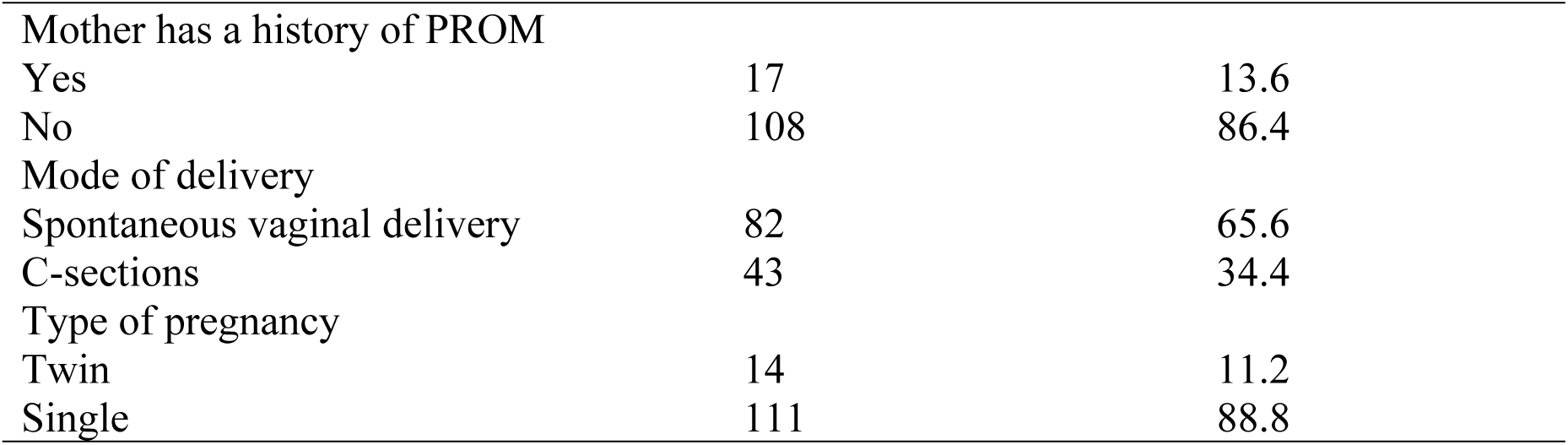
Maternal characteristics of study participants.

| Variable | Frequencies (N=125) | Percentages (%) |
| --- | --- | --- |
| Mother suffered from a bacterial infection during pregnancy |  |  |
| Yes | 81 | 64.8 |
| No | 44 | 35.2 |
| Mother had pregnancy-induced hypertension |  |  |
| Yes | 16 | 12.8 |
| No | 109 | 87.2 |
| The mother experienced an obstetric hemorrhage during pregnancy |  |  |
| Yes | 52 | 41.6 |
| No | 73 | 58.4 |
| Mother had pregnancy-induced diabetes mellitus |  |  |
| Yes | 18 | 14.4 |
| No | 107 | 85.6 |
| Mother has a history of PROM |  |  |
| Yes | 17 | 13.6 |
| No | 108 | 86.4 |
| Mode of delivery |  |  |
| Spontaneous vaginal delivery | 82 | 65.6 |
| C-sections | 43 | 34.4 |
| Type of pregnancy |  |  |
| Twin | 14 | 11.2 |
| Single | 111 | 88.8 |

### Neonatal characteristics of study participants

Regarding neonatal characteristics of study participants, 13.6% (17/125) had fever at birth, and 42.4% (53/125) had excessive secretions in the mouth and nose at birth, 15.2% (19/125) had a 5th minute APGAR score of 0–3, and 30.4% (38/125) of the babies underwent neonatal resuscitation. Furthermore, 87.2% (109/125) were term, and 19.2% (24/125) experienced fetal distress (Table 3).

**Table 3:** Neonatal characteristics.

| Variable | Frequencies (N=125) | Percentages (%) |
| --- | --- | --- |
| Fever at birth |  |  |
| Yes | 17 | 13.6 |
| No | 108 | 86.4 |
| The neonate had excessive secretions in the mouth and nose at birth |  |  |
| Yes | 53 | 42.4 |
| No | 72 | 57.6 |
| Neonate's 5<sup>th</sup> minute APGAR score |  |  |
| 0-3 | 19 | 15.2 |
| 4-7 | 63 | 50.4 |
| 8-10 | 43 | 34.4 |
| Gestational age according to birth weight |  |  |
| AGA | 83 | 66.4 |
| SGA | 16 | 12.8 |
| LGA | 26 | 20.8 |
| Baby is done with Neonatal resuscitation |  |  |
| Yes | 38 | 30.4 |
| No | 87 | 69.6 |
| Gestational age at birth |  |  |
| Term | 109 | 87.2 |
| Post-term | 16 | 12.8 |
| Weight of baby at birth(g) |  |  |
| term | 107 | 10.4 |
| Post term | 18 | 78.4 |
| Birth order of the baby |  |  |
| 2 | 52 | 41.6 |
| ≥3 | 73 | 58.4 |
| The neonate experienced fetal distress |  |  |
| Yes | 24 | 19.2 |
| No | 101 | 80.8 |

### Clinical profile of neonates with meconium-stained amniotic fluid

Tachypnoea was reported in 16.8% (21/125) of the neonates, Intercostal and subcostal retraction in 15.2% (19/125), nasal flaring in 18.4% (23/125), grunting in 9.6% (12/125), and cyanosis in 14.4% (18/125). Wheezing was reported among 10.4% (13/125) and crackles among 8.8% (11/125) (table 4).

**Table 4:** Clinical profile of neonates with meconium aspiration syndrome.

| Variable | Frequencies (N=125) | Percentages (%) |
| --- | --- | --- |
| Tachypnoea |  |  |
| No | 104 | 83.2 |
| Yes | 21 | 16.8 |
| Intercostal and subcostal retraction |  |  |
| No | 106 | 84.8 |
| Yes | 19 | 15.2 |
| Nasal flaring |  |  |
| No | 102 | 81.6 |
| Yes | 23 | 18.4 |
| Grunting |  |  |
| No | 113 | 90.4 |
| Yes | 12 | 9.6 |
| Cyanosis |  |  |
| No | 107 | 85.6 |
| Yes | 18 | 14.4 |
| Wheezing |  |  |
| Yes | 13 | 10.4 |
| No | 112 | 89.6 |
| Crackles |  |  |
| Yes | 11 | 8.8 |
| No | 114 | 91.2 |

### Radiological profile of neonates with meconium-stained amniotic fluid

Chest X-ray for MAS was positive in 6 neonates, 4.8% (6/125), bilateral infiltrates in 4.0% (5/125), hyperinflation in 5.6% (7/125), and atelectasis in 3.2% (4/125) (Table 5).

**Table 5:** Radiological profile of neonates.

| Variable | Frequencies (N=125) | Percentages (%) |
| --- | --- | --- |
| CXR for MAS |  |  |
| Negative | 119 | 95.2 |
| Positive | 6 | 4.8 |
| Bilateral infiltrates |  |  |
| Yes | 5 | 4.0 |
| No | 120 | 96.0 |
| Hyperinflation |  |  |
| Yes | 6 | 4.8 |
| No | 118 | 95.2 |
| Atelectasis |  |  |
| Yes | 4 | 3.2 |
| No | 121 | 96.8 |

### Proportion of neonates with MAS

The proportion of neonates with MAS was 4.8% (6/125), 95%CI (3.4-9.3 (**Figure 2**).

**Figure 1:**
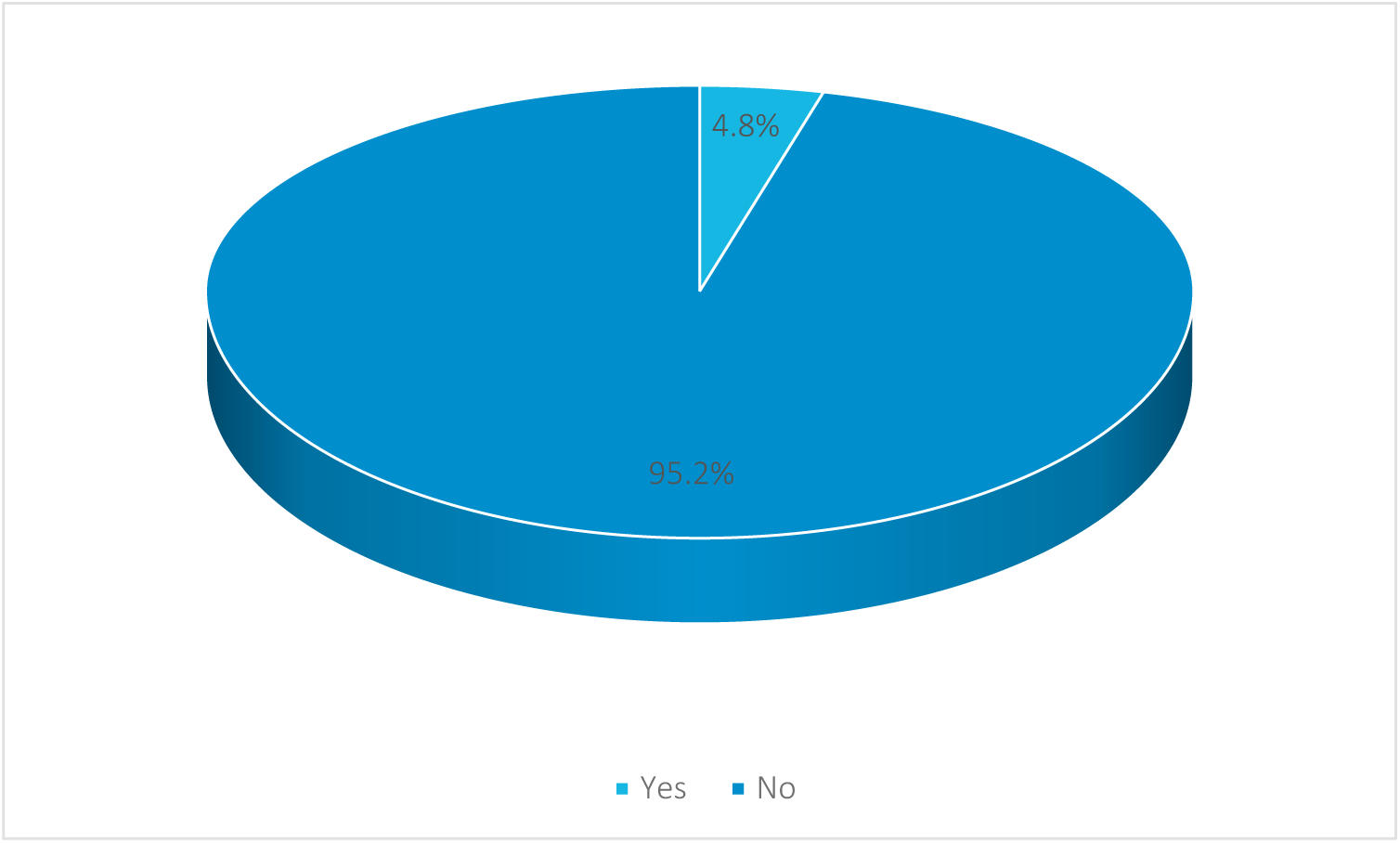
Proportion of neonates with MAS.

### Risk factors associated with MAS in neonates

#### Multivariate analysis of the factors associated with MAS among neonates

The multivariate analysis identified several independent factors significantly associated with Meconium Aspiration Syndrome (MAS) among neonates. Maternal age of 35 years and above was linked to a higher risk of MAS, with an adjusted incident rate ratio (aIRR) of 2.09 (95% CI: 1.16–13.93, *p* = 0.03). Similarly, pregnancy-induced diabetes mellitus in mothers was significantly associated with MAS, showing an aIRR of 2.77 (95% CI: 1.52–7.55, *p* = 0.04).

Among neonatal factors, fetal distress emerged as a strong predictor, with an aIRR of 3.97 (95% CI: 1.44–11.83, *p* < 0.001). Additionally, a low 5th minute APGAR score (0–3) was significantly associated with MAS, with an aIRR of 1.94 (95% CI: 1.04–6.15, *p* = 0.03) (Table 6).

**Table 6.** Multivariate analysis of the factors associated with MAS:

| Variable | Bivariate analysis |  | Multivariate analysis |  |
| --- | --- | --- | --- | --- |
|  | CIRR (95% CI) | P-value | aIRR (95% CI) | P-value |
| **Maternal age (years)** |  |  |  |  |
| <20 | Ref |  |  |  |
| 20-34 | 0.44(0.28-26.73) | 0.42 | 0.27(0.19-19.54) | 0.38 |
| 35+ | 2.32(1.07-39.51) | 0.04 | 2.09(1.16-13.93) | 0.03* |
| **Mother's education level** |  |  |  |  |
| No education | 4.09(1.78-41.83) | 0.03 | 1.87(0.91-12.83) | 0.08 |
| Primary | 1.91(0.31-31.70) | 0.49 | 1.23(0.18-8.52) | 0.21 |
| Secondary and above | Ref |  |  |  |
| **Residence type** |  |  |  |  |
| Urban | Ref |  |  |  |
| Rural | 4.47(1.93-17.41) | 0.001 | 1.73(0.93-11.82) | 0.09 |
| **Maternal factors** |  |  |  |  |
| Mother had pregnancy-induced diabetes mellitus |  |  |  |  |
| Yes | 7.50(1.78-43.82) | <0.001 | 2.77(1.52-7.55) | 0.04* |
| No | Ref |  |  |  |
| Mother has a history of PROM |  |  |  |  |
| Yes | 6.35(2.65-33.61) | 0.03 | 1.44(0.64-12.08) | 0.17 |
| No | Ref |  |  |  |
| The neonate experienced fetal distress |  |  |  |  |
| Yes | 21.04(3.78-56.61) | <0.001 | 3.97(1.44-11.83) | <0.001* |
| No | Ref |  |  |  |
| Neonate's 5<sup>th</sup> minute APGAR score |  |  |  |  |
| 0-3 | 3.39(1.09-8.14) | 0.003 | 1.94(1.04-6.15) | 0.03* |
| 4-7 | 0.34(0.14-17.50) | 0.57 | 1.13(0.45-12.85) | 0.15 |
| 8-10 | Ref |  |  |  |
| Type of pregnancy |  |  |  |  |
| Twin | 7.93(3.87-28.53) | 0.03 | 1.99(0.93-8.37) | 0.28 |
| Single | Ref |  |  |  |

*MAS=meconium aspiration syndrome, aIRR=adjusted incident rate ratio, p=p-value*

**Table 7.** Short-term outcomes among the neonates.

| Variable | No | Yes | Odds ratio | P-value |
| --- | --- | --- | --- | --- |

|  | 119(95.2%) | 6(4.8%) |  |  |
| --- | --- | --- | --- | --- |
| **In hospital deaths** |  |  |  |  |
| Yes | 2(1.7) | 4(66.7) | 117 | 0.002* |
| No | 117(98.3) | 2(33.3) |  |  |
| **Length of hospital stay** |  |  |  |  |
| >5 Days | 3(2.5) | 3(50.0) | 38.7 | <0.001* |
| ≤5 Days | 116(97.5) | 3(50.0) |  |  |

#### Short-term outcomes among the neonates

Upon Fisher’s exact test, in-hospital deaths (P = 0.002) and length of hospital stay (P < 0.001) were significantly associated with MAS among neonates in the study area (Table 6).

## Discussion

Nasal flaring was the most frequent clinical sign observed in the neonates. This finding is consistent with [10], who reported nasal flaring as a prominent symptom in neonates with MAS. Nasal flaring indicates the neonate’s effort to reduce airway resistance and improve oxygen intake. It is a sensitive early marker of respiratory distress. Its high prevalence in this study suggests that most neonates presented at a stage requiring immediate intervention, reflecting either delayed referral or inadequate antenatal risk assessment for MAS.

In this study, tachypnoea was the second most common clinical feature, observed in neonates. This finding is comparable to that of Suwera, who reported tachypnoea as one of the commonest clinical features of the neonates [11]. Tachypnoea is typically the earliest sign of respiratory distress and is due to partial airway obstruction by meconium, inflammation of the airways, and decreased lung compliance [12]. The high proportion of tachypnoea suggests a significant burden of moderate to severe respiratory compromise among affected neonates. This finding highlights the importance of prompt respiratory support at birth and close monitoring in the neonatal intensive care unit (NICU) settings, particularly in low-resource environments.

Retractions were noted in almost a sixth of neonates, indicating increased work of breathing. Similar findings were reported by [13], who observed chest wall retractions in approximately 15% of their cohort. Retractions occur as a compensatory response to negative intrathoracic pressure caused by obstructed airflow due to meconium in the airways. The presence of retractions highlights the severity of airway compromise and lung stiffness, which necessitates immediate respiratory support and may predict progression to respiratory failure if not promptly managed.

Grunting was also common among neonates. A similar frequency was reported by Mohan, who found grunting in about 8.3% of neonates with MAS [14]. Grunting is a physiological mechanism used by neonates to maintain functional residual capacity and prevent alveolar collapse. It indicates significant parenchymal lung disease and impending respiratory failure. The presence of grunting as a frequent finding implies that a majority of neonates had alveolar damage and reduced gas exchange capacity, necessitating oxygen supplementation and potentially mechanical ventilation.

Cyanosis was observed in a good number of neonates (14.5%), comparable to findings by Singh, who noted cyanosis in 13.7% of the neonates [8]. Cyanosis results from hypoxemia due to impaired oxygenation in the alveoli clogged by meconium. It is a late sign of respiratory compromise, often associated with persistent pulmonary hypertension of the newborn (PPHN). The high frequency of cyanosis in this study indicates that many neonates had progressed to advanced stages of MAS, reflecting the need for rapid oxygenation strategies and, in some cases, nitric oxide therapy where available.

Radiologically, bilateral infiltrates were seen in 4.0% of neonates, which is consistent with the findings by Ismael, who identified similar infiltrates as hallmark features of MAS [15]. These infiltrates represent widespread inflammation and atelectasis caused by chemical pneumonitis and mechanical obstruction of small airways. Their presence helps confirm the diagnosis and differentiate MAS from other causes of neonatal respiratory distress. This radiologic feature should prompt clinicians to initiate MAS-specific protocols, including respiratory support and possible surfactant therapy.

Hyperinflation was observed in 5.6% of chest X-rays. Similar findings were reported by [5], who described hyperinflated lungs due to air trapping from partial airway obstruction. Hyperinflation results in a barrel-shaped chest and reduced lung compliance. This finding is particularly important for ventilation planning, as excessive positive pressure in already hyperinflated lungs may increase the risk of barotrauma and pneumothorax. Thus, identifying hyperinflation can guide cautious ventilator settings in MAS management.

Atelectasis occurred in 3.2% of neonates. This aligns with findings by Dasgupta, who reported an incidence of pneumothorax ranging from 1.6% to 6.3% of the neonates [16]. Atelectasis may result from overdistension of alveoli due to air trapping or from aggressive resuscitation and mechanical ventilation in fragile lungs. Its presence is a severe complication that requires prompt diagnosis and intervention. The implication of this finding is the need for careful ventilatory strategies and readiness for emergency decompression in high-risk MAS cases.

### Maternal Age ≥35 Years

The study found that maternal age of 35 years and above was independently associated with a higher risk of MAS. This association may be attributed to age-related placental insufficiency, increased incidence of post-term pregnancies, and higher likelihood of comorbidities such as hypertension and diabetes, which predispose to fetal distress and meconium passage. Similar findings were reported by Singh, who observed that advanced maternal age was a significant predictor of MAS-related complications in South African neonates [8]. Clinically, this underscores the importance of intensified antenatal surveillance in older mothers, including timely induction of labour and close intrapartum monitoring to mitigate MAS risk.

### Pregnancy-Induced Diabetes Mellitus

Pregnancy-induced diabetes mellitus (GDM) was significantly associated with MAS. GDM contributes to fetal macrosomia, delayed lung maturation, and increased risk of labour complications, all of which elevate the likelihood of meconium passage and aspiration. Luo also noted that infants born to diabetic mothers had higher rates of respiratory distress and required prolonged NICU care [17]. In clinical practice, this finding highlights the need for rigorous glycaemic control during pregnancy and proactive delivery planning for diabetic mothers, including consideration of early-term delivery and readiness for neonatal resuscitation.

### Fetal Distress

Fetal distress emerged as the strongest neonatal predictor of MAS. Fetal hypoxia triggers vagal stimulation, leading to meconium passage into the amniotic fluid. If aspiration occurs, it results in airway obstruction and chemical pneumonitis. This finding aligns with Rath, who reported that fetal distress was present in over 60% of MAS cases and was associated with severe respiratory compromise [18]. Clinically, this emphasizes the critical role of continuous fatal heart rate monitoring during labour and timely obstetric interventions such as caesarean delivery to prevent aspiration events.

### Low 5th Minute APGAR Score (0–3)

A low 5th minute APGAR score was significantly associated with MAS. This score reflects poor neonatal adaptation and may indicate perinatal asphyxia, which is a known precursor to MAS. [19] similarly found that neonates with APGAR scores below 3 had higher mortality and required intensive respiratory support. In clinical settings, a low APGAR score should prompt immediate evaluation for MAS, initiation of respiratory support, and close monitoring for complications such as persistent pulmonary hypertension and neurological sequelae.

In-hospital death was significantly associated with MAS, with 66.7% of neonates diagnosed with MAS succumbing during hospitalization. This mortality rate is notably higher than the global average, which ranges between 5% and 12% [20]. A similar trend was observed in a study conducted in Thailand, where Trainak reported a MAS-related mortality rate of 7.4%, with sepsis and persistent pulmonary hypertension of the newborn (PPHN) being the leading causes of death [21]. In South Africa, Singh documented a case fatality rate of 16% among neonates with MAS, with contributing factors including low Apgar scores, PPHN, and air-leak syndromes [8].

The elevated mortality rate in the current study may reflect delayed recognition of MAS, limited access to advanced neonatal care such as mechanical ventilation and surfactant therapy, and inadequate intrapartum monitoring. Clinically, this highlights the urgent need for improved obstetric surveillance, early neonatal resuscitation protocols, and capacity building in neonatal intensive care units (NICUs) to manage MAS-related complications effectively.

The study also found a significant association between MAS and prolonged hospitalization, with half of MAS cases staying beyond five days. This aligns with findings from Rao, who reported that 71% of neonates with MAS had extended hospital stays, particularly those requiring respiratory support beyond oxygen therapy [22]. Similarly, Ismael observed a median NICU stay of 12 days among neonates with severe MAS in a resource-restricted hospital in South Africa [15].

Prolonged hospitalization in MAS cases is often driven by complications such as respiratory failure, pneumothorax, and neurological sequelae. Luo emphasized that early-term neonates with MAS had the highest prevalence of neurological damage, contributing to longer recovery periods [17]. In clinical practice, extended hospital stays not only strain healthcare resources but also increase the risk of nosocomial infections and parental anxiety. These findings advocate for early diagnosis, standardized treatment protocols, and targeted interventions to reduce the duration of hospitalization and improve neonatal outcomes

## Conclusion

The clinical profile of MAS in neonates with meconium-stained amniotic fluid at Fort Portal and Mubende regional referral hospitals were tachypnoea, intercostal and subcostal retraction, nasal flaring, grunting, cyanosis, wheezing and crackles.

The common radiological profile of neonates with meconium-stained amniotic fluid at Fort Portal and Mubende regional referral hospitals were bilateral infiltrates, positive CXR for MAS, hyperinflation and atelectasis.

The risk factors associated with meconium aspiration syndrome among neonates with meconium-stained amniotic fluid at Fort Portal and Mubende regional referral hospitals were maternal age of 35 years and above, pregnancy induced diabetes mellitus, fetal distress and low 5^th^ minute APGAR score.

## Study limitations

Being hospital-based, it did not capture MAS cases managed at lower-level health facilities or those that did not present for hospital care, introducing potential selection bias. The short follow-up period limited the ability to assess long-term neurodevelopmental outcomes, and resource constraints may have affected the availability of advanced diagnostics such as arterial blood gases or echocardiography to confirm conditions like persistent pulmonary hypertension. Lastly, some short-term outcomes might have been underreported due to limited access to neuroimaging and follow-up testing.

## Conclusion

This study provides valuable insights into the clinical and radiological profile, risk factors, and short-term outcomes of Meconium Aspiration Syndrome (MAS) in a low-resource setting. By prospectively assessing neonates across two regional referral hospitals, the findings highlight the burden of MAS and underscore the importance of early recognition, targeted management, and improved diagnostic capacity. Strengthening neonatal care and referral systems is essential to reduce MAS-related morbidity and mortality in Uganda and similar contexts.

## Declarations

**Consent for publication:** Not applicable.

## Data availability

Data can be obtained upon requested from the corresponding author (Khalif Guled Hersi

## Competing interests

The authors confirm they have no conflicts of interest.

## Sources of funding

The study received no funding.

## Authors’ contributions

**Author contribution: KGH** was the principal investigator, conceived and designed the study, collected data, analysed data and wrote the draft of the manuscript. **KB, LG and GN** supervised the work and revised the manuscript, **MN, BA, YAH, AAF, HMY, WE, BY, AMA, AKS, and TH** participated in data collection, revised the manuscript and all authors approved the final paper.

## Data Availability

Data can be obtained upon requested from the corresponding author (Khalif Guled Hersi, All relevant data are within the manuscript and its Supporting Information files.

## Acknowledgments

We extend my sincere gratitude to the administration and staff of Fort Portal and Mubende RRH for their support and cooperation during the data collection process. Our appreciation also goes to the mothers and caregivers who participated in this study for their time and willingness to provide valuable information. We are especially thankful to my research supervisors for their guidance, constructive feedback, and encouragement throughout the research process. Lastly, we acknowledge Kampala International University for the academic support and opportunity to undertake this study.

We would like to thank all participants for their valuable contributions to the study.

## Availability of data and materials

Data is provided within the manuscript or supplementary information.

## Funding

I would like to clarify that this research study was conducted without any form of external funding.

## Conflict of interest

The authors declare no conflict interests.

## Notes

### Competing Interest Statement

The authors have declared no competing interest.

### Funding Statement

The author(s) received no specific funding for this work.

### Author Declarations

Kampala International University Research and Ethics Committee with REC number 2025-921

